# Diagnostic accuracy of six commercial SARS-CoV-2 IgG/total antibody assays and identification of SARS-CoV-2 neutralizing antibodies in convalescent sera

**DOI:** 10.1101/2020.06.15.20131672

**Authors:** Annabelle Strömer, Olaf Grobe, Ruben Rose, Helmut Fickenscher, Thomas Lorentz, Andi Krumbholz

**Author notes:** these authors contributed equally (Annabelle Strömer, Olaf Grobe, Ruben Rose). Corresponding author: Andi Krumbholz, MD, Institut für Infektionsmedizin, Christian-Albrechts-Universität und, Universitätsklinikum Schleswig-Holstein, Campus Kiel, Brunswiker, Strasse 4, 24105 Kiel, Germany.

## Abstract

The reliable detection of immunoglobulin G (IgG) or total antibodies directed against the novel severe acute respiratory syndrome coronavirus type 2 (SARS-CoV-2) is important for clinical diagnostics and epidemiological studies.

Here, we compare the diagnostic accuracy of six commercially available SARS-CoV-2 IgG (Abbott SARS-CoV-2 IgG; Diasorin Liaison^®^ SARS-CoV-2 S1/2 IgG; Epitope EDI™ Novel Coronavirus COVID-19 IgG ELISA Kit; Euroimmun Anti-SARS-CoV-2 ELISA (IgG); Mikrogen *recom*Well SARS-CoV-2 IgG) or total SARS-CoV-2 antibody assays (Roche Elecsys Anti-SARS-CoV-2).

The test sensitivities were analyzed with a set of 34 sera obtained from 26 patients after PCR-confirmed SARS-CoV-2 infection and varied from 76.9% (Euroimmun) to 96.2% (Abbott). The majority of assay results were confirmed in a laboratory-developed plaque reduction neutralization test and by a SARS-CoV-2 IgG-specific line assay including measurement of generally low IgG avidities (Mikrogen *recom*Line Coronavirus IgG [Avidität], prototype).

Moreover, 100 stored sera collected during summer 2018 (N = 50) and winter season 2018/2019 (N = 50) were included to demonstrate test specificities. These varied from 96.0% (DiaSorin) to 100% (Epitope EDI™).

A subset of sera were retested with a lateral flow test (STANDARD Q COVID-19 IgM/IgG Duo) and a considerably lower sensitivity was noted.

Overall, the diagnostic accuracy of the six SARS-CoV-2 IgG/total antibody assays was good and varied from 92.9% (Euroimmun) to 98.4% (Abbott). Due to the different specificities, results of commercially available SARS-CoV-2 antibody tests should be interpreted with caution. A high proportion of antibody-positive patient sera demonstrated neutralizing capacity against SARS-CoV-2.

## Introduction

At the end of the year 2019, Chinese local health authorities reported the occurrence of a cluster of pneumonia cases in Wuhan, a megapolis in the Hubei province [1]. Shortly after, a novel beta coronavirus – which is now designated as severe acute respiratory syndrome coronavirus 2 (SARS-CoV-2) [2] - was discovered in the bronchoalveolar lavage fluid of a patient with pneumonia [3,1]. This virus emerged globally. As of June 15, 2020, the World Health Organization reported 7,761,609 SARS-CoV-2 infections and 430,241 deaths worldwide. So far, 186,461 cases and 8791 deaths have been registered for Germany (Robert Koch-Institute, data of June 15, 2020).

The diagnosis of acute coronavirus 2019 infection (COVID-19) needs the demonstration of SARS-CoV-2 ribonucleic acids in respiratory samples by real-time reverse transcription polymerase-chain reaction (real-time RT-PCR). Their specificity has been reported with 100% [4].

More recently, the measurement of immune response against SARS-CoV-2 came into the focus of clinical diagnostics, particularly by the detection of virus-specific antibodies. The use of SARS-CoV-2 antibody tests could clarify the etiology of the disease in patients who present late, after two weeks from onset of symptoms. Moreover, these tests can demonstrate the viral spread in the community and may even identify individuals who are potentially protected from re-infection by neutralizing antibodies [4].

It is believed that the majority of developed antibodies is directed against the abundant viral nucleocapsid protein while antibodies directed against the spike protein are considered more specific and may possess neutralizing capacity [4]. The diagnostic value of serological tests, however, is limited due to their potential cross-reactivity with other human coronaviruses. Furthermore, it is still unclear, how long SARS-CoV-2 antibodies will persist and whether they are protective at all [4].

In the following, we present data on the diagnostic performance of six commercially available SARS-CoV-2 IgG or total SARS-CoV-2 antibody tests. Variations in their diagnostic sensitivity and specificity were observed. The latter could lead to a considerable number of false positive results in countries with a currently low SARS-CoV-2 prevalence such as Germany. Moreover, we demonstrate SARS-CoV-2 neutralizing antibodies in a marked proportion of convalescent sera.

## Material and Methods

This retrospective study includes 37 serum samples obtained from 26 COVID-19 patients. Blood was drawn between four and 60 days (median 19 days) after a positive real-time RT-PCR result in which parts of the SARS-CoV-2 E or N genes [5] were detected in respiratory samples. Only sera from individuals that exhibited a cycle threshold (CT) ≤ 35 (median CT 26.8) in the initial real-time RT-PCR were included. These samples should contain SARS-CoV-2 IgG/total antibodies and, therefore, were considered to determine serological assay sensitivities. Seven subjects with repeated sample entries were included in the study group. Among them were two patients, each with a serum taken 0 and 9 days before the diagnosis of COVID-19.

For calculation of assay specificities, 100 archived sera collected during summer 2018 (N = 50) and during winter 2018/2019 (50) were used. In addition, two sera exhibiting a serological pattern of a recent Epstein-Barr virus (EBV) infection (EBV VCA IgG and IgM positive, EBNA-1 IgG negative, Abbott GmbH, Wiesbaden, Germany) were included to ensure that cross-reactivity is not occurring. None of these 102 sera were expected contain SARS-CoV-2 antibodies.

Test results were also used to calculate the accuracy of the assays, i.e. the proportion of correctly identified samples from all samples.

Twelve sera that were sent to us for routine SARS-CoV-2 antibody tests without a previous SARS-CoV-2 PCR result were also included to show test performance.

The Ethics committee of the Medical Faculty of the Kiel University approved the setting of this study (AZ D467/20). All sera were stored at −20°C until testing.

The testing was done with five SARS-CoV-2 IgG assays (Abbott SARS-CoV-2 IgG; DiaSorin Liaison^®^ SARS-CoV-2 S1/2 IgG, Diasorin, Dietzenbach, Germany; Epitope EDI™ Novel Coronavirus COVID-19 IgG ELISA Kit, Epitope Diagnostics, San Diego, USA; Euroimmun Anti-SARS-CoV-2 ELISA (IgG), EUROIMMUN AG, Lübeck, Germany; Mikrogen *recom*Well SARS-CoV-2 IgG, Mikrogen GmbH, Neuried, Germany) as well as one total SARS-CoV-2 antibody test (Roche Elecsys Anti-SARS-CoV-2, Roche Diagnostics, Mannheim, Germany). The recombinant antigens of these tests cover the viral nucleocapsid protein (Abbott, Epitope, Mikrogen, Roche) or the S1 domain alone (Euroimmun) or together with the S2 domain (DiaSorin) of the spike protein. All tests were conducted strictly following the recommendations of the manufacturers on an Architect (Abbott), a Liaison XL (DiaSorin), a Cobas e 411 (Roche) or for the assays of Epitope, Euroimmun and Mikrogen on the BEP 2000 system (Siemens Healthcare GmbH, Erlangen, Germany), respectively.

In general, borderline results were counted as positive. In order to allow a better comparison of the test results, the raw data were converted to relative indices according to the decision limits specified by the manufacturer. A signal/cut-off value of <1 was valued as negative and ≥1 as positive, which corresponds to a previous study [6].

A subset of sera was tested with and without avidity reagent in an IgG line assay (Mikrogen *recom*Line Coronavirus IgG [Avidität], prototype, lot LCO042001). This assay is based on the nucleocapsid proteins of the human coronaviruses 229E, NL63, OC43 and HKU1, which are used separately as antigen, and on the nucleocapsid proteins of the “classic” SARS-CoV from 2002/2003 and SARS-CoV-2. For this, a Dynablot Plus system (Mikrogen) was used. Blots were evaluated automatically with a BLOT*rix* reader and the *recom*Scan software (Mikrogen).

Exemplarily, 18 sera were retested in a rapid lateral flow assay that distinguishes SARS-CoV-2 IgG- and IgM-antibodies (STANDARD Q COVID-19 IgM/IgG Duo, SD Biosensor, Suwon-si, Republic of Korea). The manufacturer did not report the kind of epitope used in this assay.

Forty-seven sera were also tested under biosafety level 3 conditions in a plaque reduction neutralization assay (PRNT) using an own SARS-CoV-2 isolate (M16502) and Vero cells (order no. 605372, CLS Cell Lines Service GmbH, Eppelheim, Germany). The conditions were in accordance to previous reports [7,8] with slight modifications. One to two days before infection, 1.0 × 10^5^ cells were seeded per well. The 48-well plates were then incubated under standard conditions until the cells became confluent. Directly prior to the PRNT, patient sera were heat-inactivated at 56 °C for 30 minutes and then diluted from 1:10 to 1:1280 in cell-culture medium consisting of DMEM (Bio&SELL GmbH, Feucht/Nürnberg, Germany) supplemented with 3.7 g/l NaHCO_3_, 4.5 g/l glucose, 2mM L-glutamine, and 1% (v/v) Pen/Strep/Fungi Mix (Bio&SELL). A serum dilution series was made by mixing 50 µl of each dilution step with 50 µl virus suspension containing 100 plaque forming units, followed by an incubation for 1 h at 37 °C. The cells were washed with phosphate-buffered saline (PBS, Bio&SELL GmbH), inoculated with 100 μl of these virus serum dilutions and incubated for one hour at room temperature on a rocking shaker. Then, 100 µl of the cell-culture medium extended by 20 % (v/v) fetal calf serum (FCS) was added to each well to achieve a FCS concentration of 10 % (v/v). After fixation with 4 % (w/v) paraformaldehyde in PBS, the cells were stained with aqueous solution of 1 % (w/v) crystal violet and 20 % (v/v) methanol. The stained 48-well plates were photo documented. All dilution steps were tested in quadruplicates and plaque formation was compared to an untreated cell-control and a virus control. A serum dilution ≥ 1:20 that prevented the formation of plaques by 50% compared to the virus control (PRNT_50_) was considered likely to be protective.

## Results

Thirty-seven samples taken from 26 patients with a PCR-confirmed SARS-CoV-2 infection were analyzed with six SARS-CoV-2 IgG or total antibody assays. Samples #1, #7, and #9 were taken nine days before to four days after PCR and were all found to be free of SARS-CoV-2 antibodies by the assays. These samples were not considered for calculation of sensitivity but clearly demonstrated seroconversion (Figure 1). Out of the remaining 34 samples, only one serum (#20; Figure 1) which was obtained ten days after a positive RT-PCR was tested negative for SARS-CoV-2 IgG/total antibodies in the six assays. However, this sample exhibited a PRNT_50_ 1:10 to 1:20. All other samples were reactive in at least one assay (Figure 1). With respect to the sample size, assay sensitivities ranged from 76.9% (Euroimmun, 26 individuals after diagnosis of COVID-19) to 97.1% (Abbott, 34 sera including follow-up entries) (Table 1). When three samples (#32, #33, #34; Figure 1), that all exhibited isolated reactivities in the Abbott assay but did not reveal SARS-CoV-2 neutralizing capacities in the PRNT_50_, were excluded from the data set, sensitivities varied from 87.1% to 96.8%. Twenty of the other 24 COVID-19 patients developed virus neutralizing antibodies as shown by a PRNT_50_ ≥1:20 (including two samples with a PRNT_50_ 1:10 to 1:20). A comparison of S/CO values of follow-up samples #2 to #6, #26 to #27, #28 to #29, and #32 to #33 showed no clear trend towards higher values while an increase in PRNT_50_ titer was evident in the first patient (#1 to #6) (Figure 1, Supplementary material).

**Table 1:** Diagnostic performance of six commercial SARS-CoV-2 IgG and total antibody assays.

|  | sensitivity in % |  |  | specificity in % | accuracy in % |  |  |
| --- | --- | --- | --- | --- | --- | --- | --- |
|  | number of samples |  |  |  |  |  |  |
|  | 34 | 31* | 26<sup>#</sup> | 100 | 134 | 131* | 126<sup>#</sup> |
| Assay |  |  |  |  |  |  |  |
| Abbott | 97.1 | 96.8 | 96.2 | 99.0 | 98.5 | 98.5 | 98.4 |
| Diasorin | 82.4 | 90.3 | 80.8 | 96.0 | 92.5 | 94.7 | 92.9 |
| Epitope | 82.4 | 90.3 | 80.8 | 100.0 | 95.5 | 97.7 | 96.0 |
| Euroimmun | 79.4 | 87.1 | 76.9 | 97.0 | 92.5 | 94.7 | 92.9 |
| Mikrogen | 88.2 | 96.8 | 88.5 | 98.0 | 95.5 | 97.7 | 96.0 |
| Roche | 88.2 | 96.8 | 88.5 | 99.0 | 96.3 | 98.5 | 96.8 |
| *Three sera obtained from COVID-19 cases that showed isolated reactivity in the Abbott assay were excluded in this setting. |  |  |  |  |  |  |  |
| #Sera from 26 individuals after PCR-confirmed SARS-CoV-2 infection. Follow-up sera were excluded. |  |  |  |  |  |  |  |

**Figure 1:**
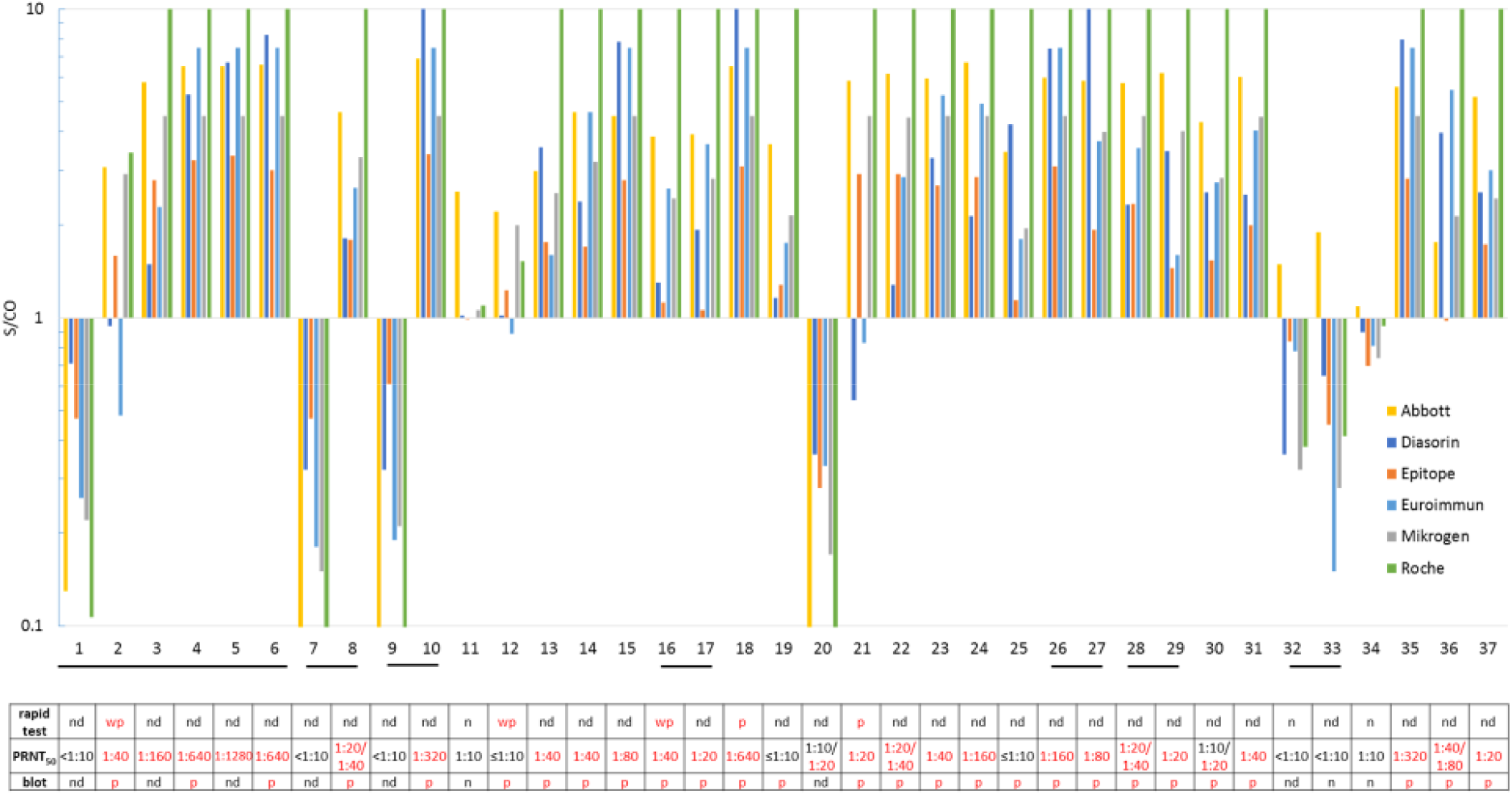
Detection of SARS-CoV-2 IgG/total antibodies in confirmed COVID-19 cases. Serum samples (N = 37) obtained from 26 COVID-19 patients were tested. A signal/cut-off value ≥1 is considered as positive for SARS-CoV-2 IgG/total antibodies. The sera marked by underlined numbers are follow-up samples from seven patients, and three among them demonstrate seroconversion. Sample #1 was taken four days and sample #2 sixth days after the PCR. The last serum (#6) of this patient was obtained 26 days after PCR. Sample #7 was taken nine days before PCR and the corresponding sample #8 19 days after the PCR. Serum #9 was taken on the day of the PCR and serum #10 29 days thereafter. Results of the rapid lateral flow test, the plaque reduction neutralization assay (PRNT_50_) and the line assay (blot) are shown in the table. P, positive; wp, weakly positive; n, negative; nd, not determined.

Specificity was calculated using 100 archived samples that should not contain SARS-CoV-2 antibodies. Overall, ten sera were found to be reactive in the six assays, and four of them were found to be reactive in one assay. None contained SARS-CoV-2 neutralizing antibodies (Figure 2, Supplementary material). Thus, assay specificities ranged from 96.0% to 100.0% (Table 1). Two samples with a typical constellation of an acute EBV infection did not show cross reactivity with any of the SARS-CoV-2 IgG/total antibody assays (data not shown).

**Figure 2:**
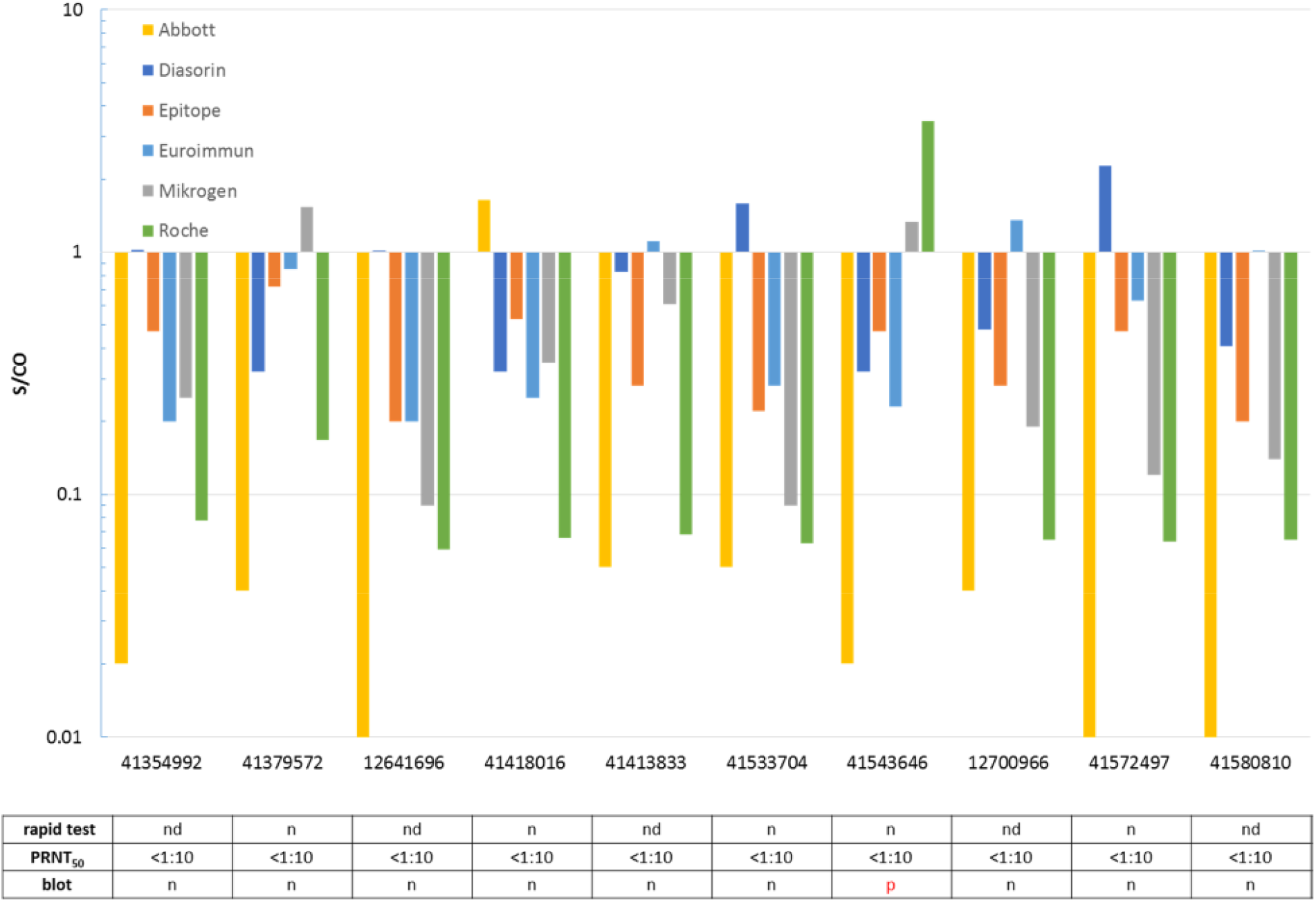
False positivity of SARS-CoV-2 IgG/total antibody tests in ten out of 100 archived sera collected in summer 2018 and winter 2018/2019. A signal/cut-off value ≥1 is considered as positive for SARS-CoV-2 IgG/total antibodies. Results of the rapid lateral flow test, the plaque reduction neutralization assay (PRNT_50_) and the line assay (blot) are shown in the table. P, positive; n, negative; nd, not determined.

Taken together, assay accuracies varied from 92.5% to 98.5% (Table 1).

Twelve routine samples obtained from individuals who were interested in their SARS-CoV-2 antibody status were re-evaluated by all six assays (Figure 3). Six of them – including three family members of a confirmed COVID-19 case (#22; Figure 1) - were classified SARS-CoV-2 IgG/total antibody positive by the majority of the tests. Isolated reactivity was observed in two sera (#2, #10; Figure 3).

**Figure 3:**
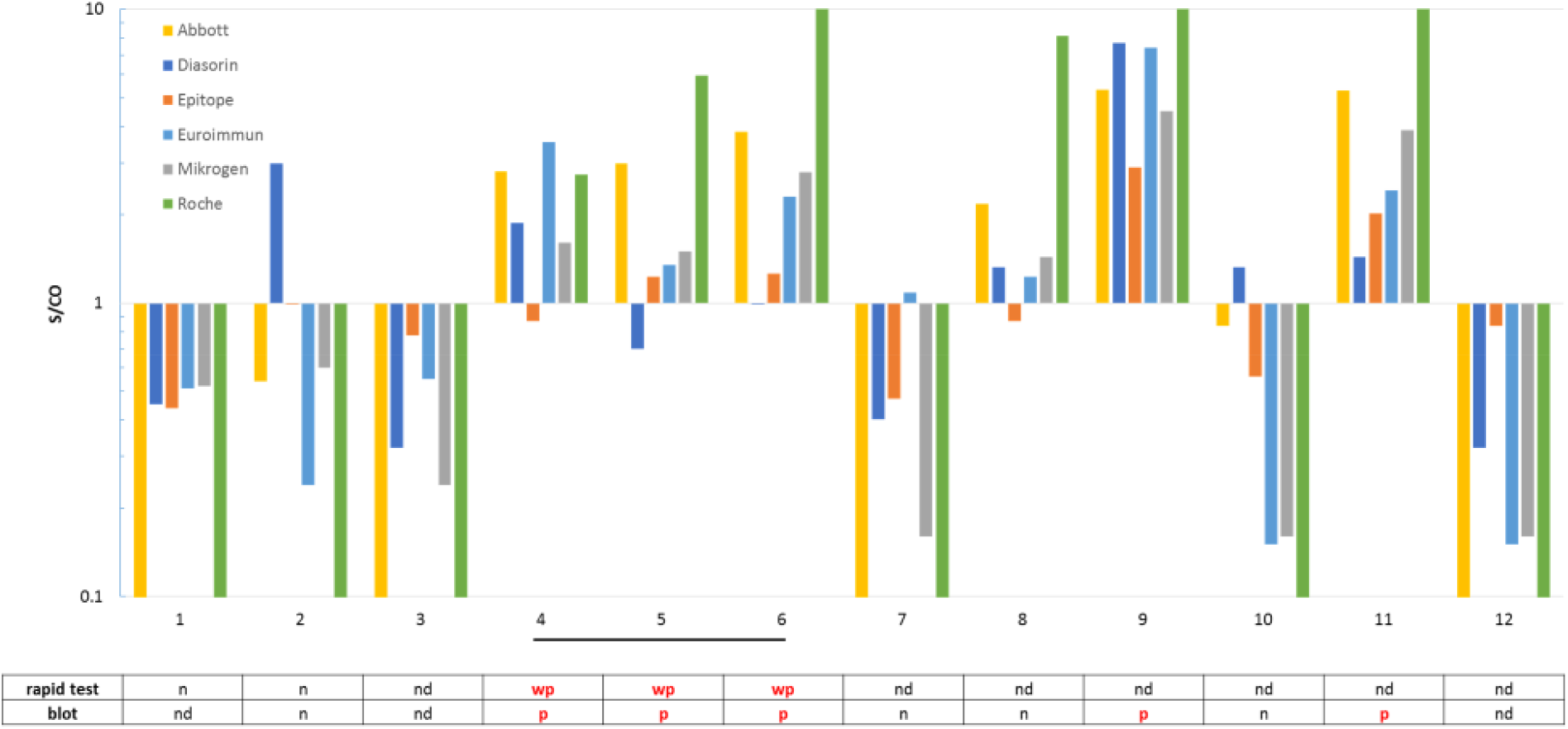
Detection of SARS-CoV-2 IgG/total antibodies in twelve routine samples. A signal/cut-off value ≥1 is considered as positive for SARS-CoV-2 IgG/total antibodies. The underlined samples #4 to #6 were obtained from family members of a COVID-19 case (sample #22 in Figure 1). Results of the rapid lateral flow test and the line assay (blot) are shown in the table. P, positive; wp, weakly positive; n, negative; nd, not determined.

Thirty of the 37 sera obtained from COVID-19 cases were retested in the IgG *recom*Line assay. Among them, 27 (90%) were confirmed as SARS-CoV-2 IgG positive (Figure 1, Supplementary material). One out of the ten archived sera that were reactive in at least one of the six SARS-CoV-2 IgG/total antibody tests was also classified as SARS-CoV-2 IgG positive by the blot results. The underlying sample was collected in winter 2018/2019 and was found to be reactive for SARS-CoV-2 IgG/total antibodies by the Mikrogen (S/SCO 1.33) and Roche (S/SCO 3.46) assays in parallel (Figure 2). As well, nine of the twelve pretested routine samples were analyzed by the blot and pre-known results were largely confirmed (Figure 3, Supplementary material). All measured SARS-CoV-2 IgGs were of low avidity (Supplementary material).

Eighteen samples, including eight sera from confirmed COVID-19 cases were retested in a lateral flow assay. Of the eight COVID-19 sera, two were clearly positive for IgG, while three were tested weakly positive and three negative (Figure 1, Supplementary). Five archived samples that were valued reactive in at least one of the six SARS-CoV-2 IgG/total antibody tests were also re-evaluated by this lateral flow assay. All of them had negative IgG results (Figure 2, Supplementary material). In addition, five routine samples were retested by the rapid antibody assay, and three were found to be weakly IgG positive (Figure 3, Supplementary material). There was no evidence of the presence of SARS-CoV-2 IgM in the 18 samples (Supplementary material).

## Discussion

In a rather short period, various assays for detection of SARS-CoV-2 antibodies were introduced to the market. A meta-analysis of 38 studies on the performance of different format SARS-CoV-2 antibody tests mainly manufactured from Chinese companies was reported recently [9]. Furthermore, studies on the sensitivity and specificity of SARS-CoV-2 IgG tests, including the ELISAs from Epitope, Euroimmun, and Mikrogen, were published previously [6,10,8,7]. Here, we compare six different commercially available tests, all of which have been released within the past few months. Three of them (Abbott, DiaSorin, Roche) were applied in a random access manner which reduces the needed hands-on-time markedly. A subset of samples were retested in a laboratory-developed PRNT_50_, in an immunoblot including the determination of SARS-CoV-2 IgG avidities and in a rapid lateral flow assay. To our knowledge, such a comprehensive study has not yet been carried out.

The sensitivities between the six tests differ from 76.9% to 97.1% (Table 1), which is in accordance to other studies [6,8,10] and to the meta-analysis [9]. Our data indicate that assays based on the more abundant viral nucleocapsid protein as an epitope are slightly more sensitive compared to tests using domains of the spike protein as an antigen. Furthermore, a lack of reactivity in the latter IgG tests does not necessarily mean that SARS-CoV-2 neutralizing antibodies are missing (compare Figure 1). However, three sera from two COVID-19 patients, which were only recognized as IgG positive in the Abbott test, could not be confirmed in the blot and in the PRNT_50_. In addition, four other COVID-19 patients did not develop a PRNT_50_ ≥1:20 (Figure 1). Thus, 76.9% (20/26) of COVID-19 patients developed SARS-CoV-2 neutralizing antibodies as demonstrated by a PRNT_50_ ≥1:20 (including two patients with a PRNT_50_ of 1:10 to 1:20). The absence of measurable neutralizing antibodies in several COVID-19 patients was reported recently [11]. The evaluability of the simple PRNT_50_ technique used in our study may be further improved by overlaying the cells with cellulose or by using specific antibodies to detect remaining viral antigens in the cells, as shown previously [7].

The specificities of SARS-CoV-2 IgG/total antibody tests are at a comparatively high level between 96.0 and 100.0%, as has also been reported by others [6,8,10]. Interestingly, one sample collected in winter 2018/2019 was found to be reactive in the Mikrogen and Roche assays as well as in the blot but could not be confirmed by the PRNT_50_ (Figure 2, Supplementary material). For all other 99 archived samples, a random pattern of rare, isolated reactivity was demonstrated and none was confirmed as possessing SARS-CoV-2 neutralizing antibodies by PRNT_50_ (Figure 2, Supplementary material). In addition, two sera obtained from patients with acute EBV infection were tested negative for SARS-CoV-2 IgG/total antibodies. Thus, cross-reactivity to epitopes of endemic human coronaviruses or triggered by active EBV infection may not represent a major problem. Nevertheless, it should be noted that due to the currently estimated low prevalence of SARS-CoV-2 antibodies in the overall German population, even a rather high specificity of 99% would produce a relevant number of false positive results. Thus, these assays - that all show a comparable high accuracy of 92.5%-98.5% (Table 1) - should be preferentially used for testing of patients with a history of a probable infection. In case of doubt, the implementation of the labor-intensive and time-consuming PRNT_50_ should be considered.

The majority of SARS-CoV-2 IgGs in sera from COVID-19 patients were confirmed by an immunoblot but were shown to possess a low avidity (Supplementary material). This result may come from the short observation period. The last sample was obtained 60 days after COVID-19 diagnosis but the median of sample collection dates was only 19 days after PCR. Furthermore, seroconversion in three SARS-CoV-2 patients was demonstrated. So far, it is not clear when SARS-CoV-2 IgGs of high avidity will appear and how long such SARS-CoV-2 specific antibodies will persist at all. A marked proportion of COVID-19 patients, however, developed neutralizing antibodies in their sera, which might be protective and prevent a re-infection.

Lateral flow assays for the detection of SARS-CoV-2 antibodies are becoming increasingly important as they can be used by less experienced persons under point-of-care conditions and should lead to reliable results in a relatively short time [8]. Here, 18 sera were retested with a rapid antibody test. Only two convalescence sera obtained from COVID-19 patients were clearly identified as IgG positive by this assay. In addition, only very weak bands were visible in three of these well-characterized samples, while three further convalescent sera were rated negative (Supplementary material). A similar situation with hardly visible IgG bands was observed in three routine samples obtained from family members of a confirmed SARS-CoV-2 case (Supplementary material). In contrast, five archived samples that were classified reactive for SARS-CoV-2 IgG/total antibodies by at least one of the six assays were all negative by this lateral flow assay. Thus, this rapid antibody test is believed to have a good specificity but sensitivity is reduced. Particularly the occurrence of faint IgG bands is a diagnostic challenge. The interpretation of results obtained from this assay should be done with caution.

Taken together, all six tested SARS-CoV-2 IgG/total antibody assays demonstrate a good performance. Their slightly reduced specificities, however, may produce a relevant number of false positive results in low prevalence countries. Future studies should address the long-term course of SARS-CoV-2 antibodies as well as the virus-specific cellular immune response. For the latter, the development of routine test procedures is urgently required. The PRNT results indicate that a high proportion of antibody-positive sera are actually able to neutralize the virus in culture and, thus, may be relevant for immunity.

## Data Availability

The data are available in the supplement.

## Acknowledgments

The authors deeply acknowledge the excellent technical assistance of Sina Müller (Labor Dr. Krause und Kollegen MVZ GmbH) and Andrea Hölzgen (Institut für Infektionsmedizin). Furthermore, we would like to thank all patients for participating in this study.

## Author’s contributions

AK conceived the study and supervised all experiments. AS and OG prepared samples. AS performed testing of all sera in the commercially available assays with help of OG. RR performed the PRNT with support by AK and HF. AK, AS, OG and RR prepared the manuscript with help of HF and TL. Results are part of the MD thesis of AS.

